# Insomnia-Related Metabolomic Profiles Reflect Antioxidant Deficits and Relate to Cognitive Decline Through a Metabolic Risk Score in HCHS/SOL

**DOI:** 10.64898/2026.02.04.26345594

**Authors:** Cynthia Kusters, Jose Santos Cabrera, Yu Zhang, Ying Zhang, Tianyi Huang, Joon Chung, Bing Yu, Qibin Qi, Carmela Alcantara, Wassim Tarraf, Krista M Perreira, Raanan Arens, Alberto R. Ramos, Martha L. Daviglus, Phyllis C. Zee, Hector M. González, Carmen R. Isasi, Susan Redline, Tamar Sofer

**Affiliations:** Department of Epidemiology, Fielding School of Public Health, University of California, Los Angeles, Los Angeles, CA, USA; Department of Epidemiology, Harvard T.H. Chan School of Public Health, Boston, MA, USA; Division of Sleep Medicine and Circadian Disorders, Department of Medicine, Brigham and Women’s Hospital, Boston, MA, USA; Channing Division of Network Medicine, Brigham and Women’s Hospital, Harvard Medical, Boston, MA, USA; CardioVascular Institute (CVI), Beth Israel Deaconess Medical Center, Boston, MA, USA; Laboratory of Epidemiology and Population Sciences, Intramural Research Program, National Institute on Aging, Baltimore, MD, USA; Department of Medicine, Harvard Medical School, Boston, MA, USA; Department of Epidemiology, School of Public Health, The University of Texas Health Science Center at Houston, Houston, TX, USA; Department of Epidemiology and Population Health, Albert Einstein College of Medicine, Bronx, NY, USA; School of Social Work, Columbia University, New York, NY, USA; Institute of Gerontology, Wayne State University, Detroit, MI, USA; Department of Social Medicine, University of North Carolina, Chapel Hill, North Carolina, USA; Division of Pediatric Respiratory and Sleep Medicine, Department of Pediatrics, Children’s Hospital at Montefiore, Bronx, New York, NY, USA; Sleep Medicine Program, Department of Neurology, University of Miami Miller School of Medicine, Miami, FL, USA; Institute for Minority Health Research, University of Illinois, Chicago, IL, USA; Division of Sleep Medicine, Department of Neurology, Northwestern University, Chicago, IL, USA; Department of Neurosciences and Shiley-Marcos Alzheimer’s Disease Center, University of California, San Diego, La Jolla, CA, USA; Department of Biostatistics, Harvard T.H. Chan School of Public Health, Boston, MA, USA

**Author notes:** These authors contributed equally to this work (Cynthia Kusters, Jose Santos Cabrera). Corresponding authors: Cynthia Kusters, 650 Charles E. Young Dr. South, Center for Health Sciences, Los Angeles, CA 90095-1772, and Tamar Sofer, Center for Life Sciences CLS-934, 3 Blackfan St, Boston, MA 02115.

**Keywords:** Untargeted metabolomics, insomnia, Hispanic/Latino adults in the United States, discovery and replication

## Abstract

**Background:** We aimed to identify metabolites and create risk scores for insomnia symptoms in U.S. Hispanic/Latino adults.

**Methods:** We analyzed data from 6,107 participants in the Hispanic Community Health Study/Study of Latinos, split into discovery (n=3,932) and replication datasets (n=2,175). Serum metabolites and the Women’s Health Initiative Insomnia Rating Scale (WHIIRS) were collected at baseline. We examined the relationships between 768 metabolites and insomnia symptoms and suspected insomnia (WHIIRS≥9) using the discovery dataset, followed by replication. Metabolite risk scores (MRSs) were generated with LASSO regression and evaluated for replication. We assessed the relationships of replicated metabolite measures and MRS with sleep, cognitive, and psychological traits (cross-phenotypes).

**Findings:** Nine metabolites were associated with insomnia symptoms in the discovery study, with two of these being replicated. Lower levels of hydrocinnamate and indolepropionate correlated with increased insomnia symptoms. We developed MRS for insomnia symptoms with replication. Various associations were observed between the two metabolites, 2 MRS, and cross-phenotypes. For instance, the WHIIRS MRS was associated with a higher risk of mild cognitive impairment (MCI) seven years later (OR:1.58, 95%CI:1.43-1.74 per 1 SD increase in MRS).

**Interpretation:** The metabolomic profile associated with insomnia symptoms encompasses diet and gut microbiome metabolites. This study identified specific metabolites linked to insomnia that are also related to comorbidities, such as a higher risk of developing MCI during follow-up, suggesting a shared mechanism.

**Funding:** Grants from various National Institutes of Health and the JLH Foundation supported the work.

**Research in context:** *Evidence before this study:* Insomnia affects 30–36% of individuals, with clinical insomnia estimated at 6–10%, and it is more severe among Hispanics, who also face higher risks for cognitive decline and cardiovascular disease. While previous metabolomics studies have investigated sleep disorders, most have focused on sleep apnea or sleep duration, not insomnia. The few studies that focus on insomnia were limited by small sample sizes or co-occurring psychiatric conditions. Only two large-scale studies linked insomnia symptoms to specific metabolites, but neither examined these associations in Hispanics or their connection to cognitive decline—gaps this study aims to address using data from the HCHS/SOL cohort.

*Added value of this study:* We identified nine metabolites related to insomnia symptoms, with two—hydrocinnamate and indolepropionate—being replicated. We also created and validated metabolite risk scores (MRS), which predicted a higher likelihood of developing mild cognitive impairment (MCI) seven years later. These results provide new insights into the metabolic pathways connecting insomnia and cognitive decline in a high-risk Hispanic population.

*Implications of all the available evidence:* Our findings indicate that insomnia symptoms are linked to specific metabolic changes, some of which may also play a role in cognitive decline. Identifying metabolites related to diet and the gut microbiome points to biological pathways that could be modified through lifestyle or therapeutic interventions. The metabolite risk scores (MRS) developed in this study showed links with mild cognitive impairment (MCI) over time, suggesting their potential usefulness in understanding long-term health risks associated with sleep disturbances. These results encourage further research into the role of metabolomics in sleep and cognitive health, especially in high-risk populations like Hispanics.

## Introduction

Insomnia, defined as difficulty initiating, maintaining sleep or early morning awakening accompanied by daytime dysfunction,[1] is a common condition known to have a direct negative impact on quality of life.[2] The prevalence of this sleep disorder varies between 5 and 50%, depending on the criteria used and the study population. On average, approximately 30-36% of individuals report experiencing at least one symptom of insomnia. When applying more stringent clinical criteria, the estimated prevalence of insomnia is around 6-10%.[3] Insomnia symptoms increase with age, and have been reported to be more severe among Hispanics compared to non-Hispanic whites.[4]

Individuals with insomnia are known to be at increased risk for developing or experiencing many health-related outcomes, such as an increased risk of hypertension,[5] cardiovascular disease,[5,6] poor cognitive performance[7,8] and, prospectively, with an increased risk of dementia.[9] Notably, this group exhibits a higher prevalence of uncontrolled hypertension, even though the overall rate of hypertension is comparable to other demographics.[10] Additionally, research indicates that the Hispanic population experiences elevated rates of cognitive impairment and dementia,[11] alongside a higher prevalence of major cardiovascular disease risk factors.[12] As insomnia impacts these risks, it is essential to identify the molecular basis or pathways involved. This project aims to explore this question through the analysis of plasma metabolomics.

Within untargeted metabolomics analysis, we assess the association between numerous metabolites derived from mass spectrometry or nuclear magnetic resonance spectroscopy from biofluids, most commonly blood plasma or serum.[13,14] This method not only explains mechanisms but also has the potential to derive potential biomarkers for exposure or disease status, offering a promising avenue for future research and clinical applications. [15] Previous metabolomics studies have tried to identify metabolites associated with sleep disorders,[16–26] with most studies focusing on obstructive sleep disorders and sleep duration. As reviewed in Humer et al,[19] there is comparatively less research on insomnia or its symptoms in the context of metabolomics.[27–30] The inference we can make based on these studies is limited as they were either based on small sample sizes or reviewed specific clinical populations with additional symptoms of depression and anxiety that may skew the results. At this point, only two extensive studies have examined the association between insomnia or insomnia-related symptoms, such as poor sleep quality, and metabolomics.[28] A study of predominantly white, non-Hispanic/Latino, middle-aged, and older women from the Women’s Health Initiative and Nurses’ Health Studies demonstrated that women reporting poor sleep quality had elevated shorter and more saturated lipids. [28] In another extensive study, based on various Dutch cohorts, insomnia symptoms were associated with lower citrate, higher glycoprotein acetyls, and lower phospholipids in very large HDL particles. These findings were verified using a Mendelian randomization analysis using data from the UK Biobank.[31] In addition to identifying individual metabolites and corresponding pathways, metabolomics analysis can also develop metabolic scores that can serve as biomarkers for the underlying mechanisms and as potential predictors of future outcomes.[32] Previous metabolic scores for sleep-related outcomes have successfully linked these specific sleep-related scores to coronary heart disease,[28] hypertension,[33], obesity,[34] and diabetes.[33,34] However, although insomnia has been linked to cognitive decline, no study has examined the relationship between metabolic profile changes associated with insomnia symptoms and risks for cognitive decline. With an aging population and a rising prevalence of cognitive decline, this study aims to identify individual metabolites associated with insomnia symptoms in a cross-sectional analysis among a population, specifically Hispanics from the Hispanic Community Health Study/Study of Latinos (HCHS/SOL), who are at increased risk for both insomnia and cognitive decline. Additionally, we seek to develop an overall metabolite score and examine its prospective relationship with cognitive decline.

## Methods

To investigate the associations between metabolomics and insomnia in U.S. Hispanic/Latino adults, we conducted analyses as illustrated in Figure 1. Briefly, this study is divided into two primary analyses, followed by further examination of the associations found in the primary analyses, cross-phenotype analysis, and additional sensitivity analyses to ensure the validity of our findings. The two primary analyses performed were a metabolite-wide association study (MWAS) for both continuous measures of insomnia symptoms and a dichotomized estimate of suspected insomnia diagnosis (step 1). Second, we developed a metabolite risk score for suspected insomnia and insomnia symptoms (step 2). Both primary analyses were conducted on a discovery subpopulation, followed by replication within a second independent study set. Individuals in this study were from the Hispanic Community Health Study/Study of Latinos (HCHS/SOL).

**Figure 1.**
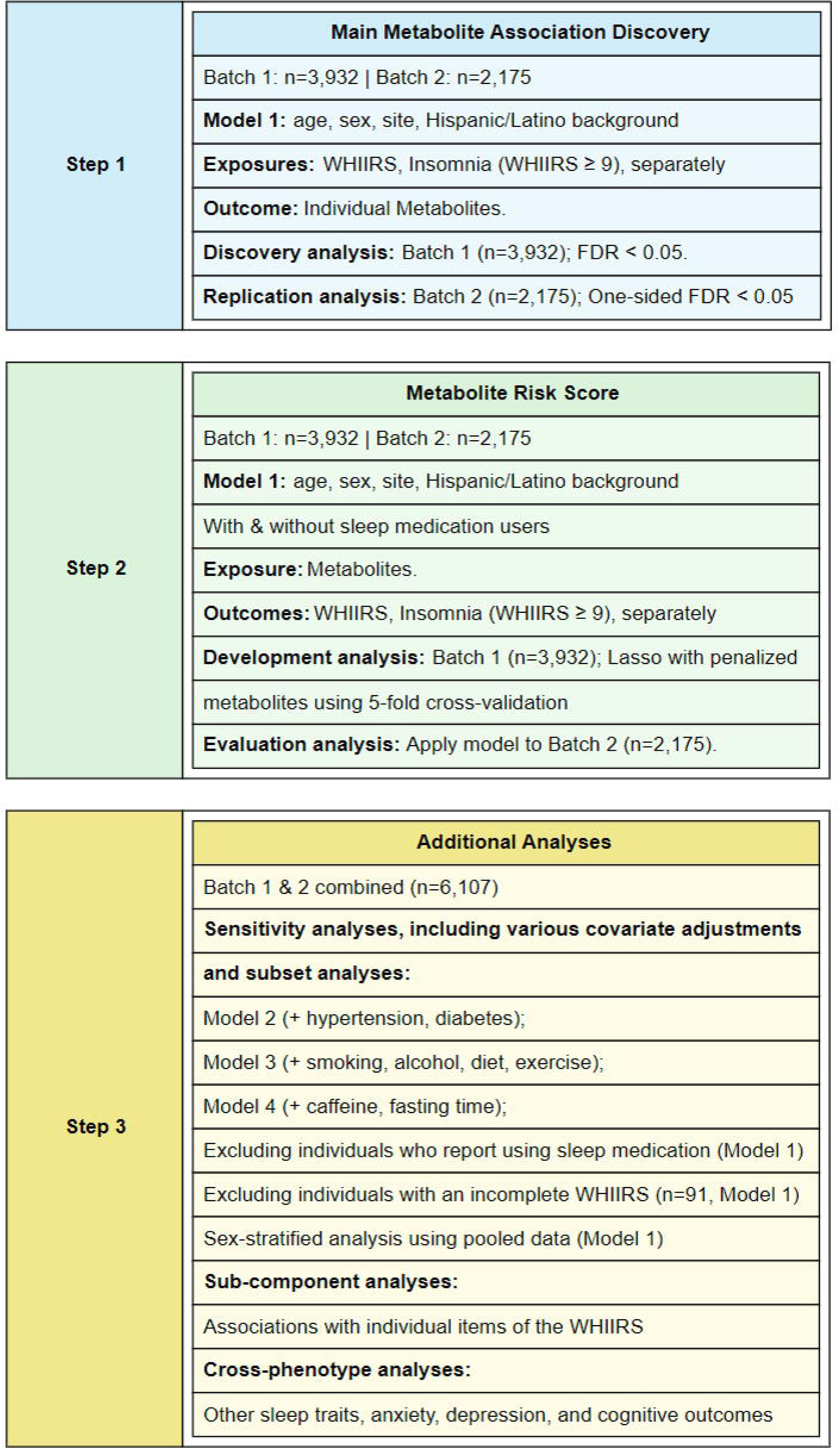
Flowchart of analyses studying metabolite associations with insomnia. The primary analysis performed single-metabolite association analyses of WHIIRS and suspected insomnia, with a discovery-replication approach using distinct HCHS/SOL datasets (step 1). We also developed metabolite risk scores for both WHIIRS and suspected insomnia, as seen in step 2. We then performed additional analyses on the combined pooled dataset (batches 1 and 2), as shown in step 3. These can be divided into three categories: 1) Sensitivity analyses with various covariate adjustments and subset analyses (excluding medication users and sex-stratified). 2) Sub-component analyses for the individual items of the WHIIRS. And 3) Cross-phenotype analysis, where we analyzed the associations with other sleep phenotypes, anxiety, depression, and various cognitive outcomes. The analyses were adjusted for the same variables as in Model 1, unless otherwise specified in the figure.

### The Hispanic Community Health Study/Study of Latinos (HCHS/SOL)

HCHS/SOL is a multicenter, longitudinal cohort study following self-reported Hispanic/Latino individuals in the U.S. aged 18 to 74 years. Participants were recruited [32,33] from four geographic areas: the Bronx, NY; Miami, FL; Chicago, IL; and San Diego, CA [32,33]. The sample design and implementation of probability sampling weights have been described in detail in previous publications [32,33], which utilized a complex study design, including stratified, two-stage sampling [31,32]. Probability sampling weights, adjusted for non-response, are provided by the HCHS/SOL Data Coordinating Center (DCC) to facilitate descriptive statistics and parameter estimates that apply to the HCHS/SOL target population.

For this study, we utilized data from the HCHS/SOL study’s baseline exam, including both phenotypic data (such as sleep-related measurements) and metabolomics data derived from blood samples. Additionally, for the cognitive assessment, we used follow-up data from participants during their second HCHS/SOL visit as part of the Study of Latinos-Investigation of Neurocognitive Aging (SOL-INCA) ancillary study, approximately seven years after their first visit.

### Baseline phenotypic data

The study’s baseline exam, conducted between 2008 and 2011, collected a range of data, including clinical assessments and questionnaires. These included extensive background information on lifestyle, medical history, social, economic, and psychosocial characteristics, as well as cognitive functioning, clinical measurements of blood pressure, anthropometrics, and electrocardiography, and laboratory measures, including lipids, blood counts, kidney function, and glycemic traits.[35] Sleep-related information is described in more detail below.

### Insomnia symptoms

Insomnia symptoms at the baseline HCHS/SOL exam were assessed with the Women’s Health Initiative Insomnia Rating Scale (WHIIRS). The WHIIRS is well validated across various race/ethnic groups (including Hispanics),[34–36] and suspected insomnia (binary) was defined as WHIIRS≥9 and “no insomnia” was defined by WHIIRS<9.[36–38] The WHIIRS is based on five questions, each with a Likert scale ranging from 0 to 4, referring to the last 4 weeks: trouble falling asleep, difficulty waking up, frequent awakenings throughout the night, and a question about overall sleep quality. A small subset of participants had one or two missing responses on the WHIIRS (n=88 and n=3, respectively); for these individuals, we imputed the missing values using the mean of their completed responses. Our primary outcomes in this study were the continuous WHIIRS scores and dichotomized suspected insomnia based on the WHIIRS threshold.

### Metabolomics data

Metabolomics measures were collected from baseline blood samples as described below. The blood samples were collected for two independent ancillary sub-studies from the HCHS/SOL, enabling separate discovery and replication analyses —the gold standard for verifying associations using high-dimensional data, such as metabolomics. The first metabolomics ancillary study (batch 1) assayed metabolomics in 4,002 HCHS/SOL participants randomly selected from those with genetic data in 2017.[39,40] The second set of metabolomics assays, collected in 2021 and referred to as batch 2, was obtained from 2,178 individuals with stored blood samples that met the following criteria. First, the sampling included specific subsets of individuals, such as those who participated in the ECHO-SOL ancillary echocardiogram study,[41] and those with a normal estimated glomerular filtration rate (eGFR>69) at the baseline HCHS/SOL exam experienced a substantial decline by the second HCHS/SOL exam. Second, the sampling also included a random selection of individuals with eGFR measurements available from both the baseline and second HCHS/SOL exams. Third, it included over 50 samples for quality control analysis between the first and second metabolomics batches; these overlapping samples were excluded from the second batch for our analysis to ensure independence between the discovery and replication sets.

A comprehensive description of the metabolomic assay protocol is provided in Supplementary Note 1. In brief, serum samples were collected after fasting at the baseline exam. Metabolomics assays were performed by Metabolon Inc. (Durham, NC) using their Discovery HD4 platform. As described in Supplementary Figure 1 (sample and metabolite selection for analysis), batch 2 included duplicated samples, of which we randomly selected one sample per individual (samples from 38 individuals). In addition, we excluded samples from batch 2 if corresponding samples were already included in batch 1 (n=128). Metabolites with more than 25% missing values across individuals per batch were excluded. Of the remaining metabolites, we replaced missing values with half the minimum observed value across individuals per batch. Finally, each metabolite’s values were rank-normalized across individuals within each batch. In the pooled analyses, the two datasets were aggregated after normalization.

After initial quality control across the metabolomics batches, the dataset included 3,978 individuals in batch 1 and 2,202 in batch 2 (see Supplementary Figure 1). Following excluding individuals with missing WHIIRS data, 3,932 individuals remained for the insomnia-metabolite association analysis in batch 1 and 2,175 in batch 2. The metabolomics data set after quality control included 768 metabolites that were available in both batches.

### Cross-phenotype assessments

#### Other sleep and psychological assessments at baseline

Other sleep measures used in our cross-phenotype analyses included time in bed, based on self-reported times the participant would go to bed and wake up on both weekdays and weekend days, from which we estimated sleep duration as the weighted average of weekday and weekend sleep. Short and long sleep were defined as time in bed ≤ 6 hours and >9 hours, respectively. Approximately 90% of the participants had a standardized sleep test where the respiratory event index was measured (REI).[42] For this study, we dichotomized participants into mild-to-severe obstructive sleep apnea (OSA, REI>5) versus no indication of OSA (REI≤5). Information about an individual’s work schedule was self-reported as day, afternoon, night, shift, irregular, and rotating shift. For this analysis, we will categorize individuals as either shift workers or day workers, with anyone reporting non-day work classified as a shift worker.

Psychological outcomes were anxiety symptoms measured by the Spielberger Trait Anxiety Inventory scale (STAI-10),[43] and depression symptoms estimated by the Center for Epidemiological Studies Depression scale (CESD-10).[44] For suspected depression, we applied a CESD-10 threshold of 10 or higher, and for an elevated anxiety status, we utilized a STAI-10 threshold of more than 20.[45]

### Cognitive assessments at baseline and follow-up

For cross-phenotype assessments, we used several cognitive outcomes. During the baseline HCHS/SOL exam, neurocognitive tests were administered to 9,714 participants who were 45 years old, consented, and had no health limitations for the neurocognitive testing. Then, during the second visit, on average 7 years after baseline, 6,377 participants completed additional neurocognitive tests during the SOL-INCA ancillary study.[46–48] For the cross-phenotype assessments, we used the following cognitive outcomes: (1) global cognitive function at baseline, (2) global cognitive change from baseline to the SOL-INCA exam (the difference between the score at follow-up minus the score at baseline), and (3) mild cognitive impairment (MCI) status, which included the classification of “MCI+” (more severe MCI, which could represent dementia) evaluated at the follow-up exam. These outcomes were described in more detail earlier.[49,50] Global cognitive function was assessed as the average of z-scores from four cognitive tests evaluating verbal episodic learning and memory, word fluency, processing speed, and executive function. These tests were administered at both the baseline and follow-up exams. Scores were calculated only for individuals with complete data for all cognitive tests. MCI status was defined based on the National Institute on Aging-Alzheimer’s Association (NIA-AA) criteria.[51] MCI status was determined for 3,149 participants with metabolomics data and available WHIIRS scores.

### Statistical analysis

#### Metabolite-wide association analysis (MWAS)

We performed a survey, linear and logistic regression analysis to assess the association between metabolites and WHIIRS and suspected insomnia in batch 1, respectively. In the model, we designated metabolites as the outcome and the insomnia measure as the exposure. For the primary analysis, we adjusted for age, sex, study center, and self-reported Hispanic/Latino background (also referred to as Model 1 throughout this manuscript). Statistically significant associations were defined according to Benjamini-Hochberg False Discovery Rate (FDR)[52] adjusted p-values less than 0.05. These metabolites were subsequently tested in the replication study (batch 2). We considered them replicated if the FDR-adjusted one-sided p-value was less than 0.05. We used FDR-adjusted one-sided p-values to confirm replication only when the direction of association was consistent between discovery and replication batches.[53]

### Metabolite risk score for insomnia

We developed a metabolite risk score (MRS) for suspected insomnia and insomnia symptoms using the same methodology applied to sleep-disordered breathing.[20,33] Specifically, we applied LASSO penalized regression without incorporating survey weights using the glmnet R package (version 4.1-8). We conducted separate regression analyses for suspected insomnia and insomnia symptoms using data from batch 1. Each model included unpenalized covariates and all potential penalized metabolites. Metabolite selection was performed using LASSO regression, with the tuning parameter (*λ*) derived from a 5-fold cross-validation. Based on the selected metabolites and their corresponding LASSO coefficients, we calculated the MRS as a weighted sum of the rank-normalized metabolite values in batch 2. The MRS was validated in batch 2 by assessing its association with suspected insomnia and insomnia symptoms in logistic and linear survey regression, adjusted for the same covariates used in its development. To account for potential confounding by sleep medication use, we also created the MRS after excluding individuals who reported using any sleep medication.

### Sensitivity analysis

To ensure the robustness of our findings, we performed additional sensitivity analyses for our MWAS analyses. We pooled the data from batches 1 and 2 for these sensitivity analyses. As a first sensitivity analysis, we modified the covariate set by adding additional potential confounders to assess the impact of residual confounding on the association between insomnia measures and metabolites. Our original model was adjusted for age, sex, study center, and self-reported Hispanic/Latino background. We used three additional regression models, with Model 1 as the primary model and the other models used to evaluate the potential impact on the insomnia measures-metabolite associations by potential confounders. Model 2 included the same covariates as Model 1 and added two cardiometabolic disease indicators: hypertension and diabetes mellitus; and Model 3 additionally included three lifestyle measures: smoking status (current, former, never smoker), alcohol drinking status (current, former, never), and the Alternative Health Eating Index (AHEI, 2010 version);[54] Model 4 added the fasting time and daily caffeine intake as potential confounders (49)). We conducted additional sensitivity analyses [51] using the covariates from our primary analysis while excluding individuals who reported any use of sleep medication. We also performed our primary analysis excluding individuals with missing component questions of the WHIIRS (n=91). Finally, we performed a sex-stratified analysis to assess potential heterogeneity in our findings.

### Subcomponent analysis

For the metabolites reported as associated with suspected insomnia or WHIIRS in the primary MWAS analysis and replicated in batch 2, as well as the MRS for suspected insomnia and WHIIRS, we conducted subcomponent analysis of the individual WHIIRS insomnia symptom questions. We also developed MRSs for each questionnaire item used in the WHIIRS to compare overlap patterns between the metabolites selected by LASSO.

### Cross-phenotype analysis

In addition to the subcomponent analysis, we examined the association between these metabolites and MRSs and co-morbid sleep phenotypes. Specifically, we assessed the association with short time in bed (short sleep), long time in bed (long sleep), mild-to-severe OSA, and shift work. Furthermore, we evaluated the association with psychological and cognitive functioning, using depression symptoms as a CESD10 score of 10 or higher (indicative of suspected depression), anxiety symptoms as a STAI10 score of 20 or higher (indicative of elevated anxiety status), as well as global cognitive function at baseline, global cognitive change from baseline to follow-up, and MCI at the follow-up visit. To increase statistical power, particularly for cognitive outcomes measured in a subset of individuals, we performed these analyses using a pooled dataset from batches 1 and 2.

### Ethics statement

The HCHS/SOL was approved by the institutional review boards (IRBs) at each field center, where all participants gave written informed consent, and by the Non-Biomedical IRB at the University of North Carolina at Chapel Hill, to the HCHS/SOL Data Coordinating Center. All methods and analyses of HCHS/SOL participants’ materials and data were carried out in accordance with human subject research guidelines and regulations.

## Results

Table 1 characterizes the HCHS/SOL study population stratified by batch, whereas supplementary Table 1 provides characteristics stratified by sex. Several characteristics differed between the two batches. For example, in batch 2, there were more female individuals (57% vs. 51%), their average age was older (53 vs. 46 years), fewer current smokers (18% vs. 25%) and fewer alcohol drinkers (42% vs. 54%). In accordance with the older age, the prevalence of diabetes (26% vs. 15%), mild-to-severe OSA (37% vs. 27%), and suspected insomnia (37% vs. 33%) was higher. The older age and higher prevalence of females in batch 2 could explain the higher proportion of suspected insomnia.

**Table 1:** Participant characteristics in the two batches with metabolomics and WHIIRS (n=6,107)

|  |  |  |  |
| --- | --- | --- | --- |
|  |  | **Batch 1** | **Batch 2** |
| N |  | 3,932 | 2,175 |
| Gender (%) | Female | 2,248 (50.7) | 1,410 (57.1) |
| Center (%) | Bronx | 1,016 (28.4) | 558 (27.6) |
|  | Chicago | 909 (14.7) | 551 (14.4) |
|  | Miami | 1,115 (32.2) | 616 (38.3) |
|  | San Diego | 892 (24.7) | 450 (19.7) |
| BMI (mean (SD)) |  | 29.78 (6.30) | 30.20 (5.80) |
| Age (mean (SD)) |  | 45.86 (15.14) | 52.70 (13.18) |
| Background (%) | Dominican | 377 (10.2) | 274 (12.8) |
|  | Central American | 399 (6.6) | 232 (6.7) |
|  | Cuban | 668 (22.7) | 405 (30.4) |
|  | Mexican | 1,430 (34.7) | 671 (27.1) |
|  | Puerto Rican | 696 (16.8) | 387 (16.1) |
|  | South American | 231 (4.5) | 174 (5.6) |
|  | More than one/Other heritage | 123 (4.4) | 30 (1.3) |
| Alcohol use (%) | Never | 727 (16.7) | 538 (26.1) |
|  | Former | 1,259 (28.8) | 721 (31.7) |
|  | Current | 1,945 (54.4) | 914 (42.2) |
| Smoking (%) | Never | 2,297 (59.4) | 1,282 (59.7) |
|  | Former | 787 (16.0) | 495 (22.4) |
|  | Current | 845 (24.5) | 396 (17.8) |
| Diabetes (%) | Yes | 738 (15.0) | 610 (26.1) |
| Hypertension (%) | Yes | 1,124 (23.0) | 849 (37.9) |
| AHEI 2010 (mean (SD)) |  | 48.57 (7.52) | 49.91 (7.37) |
| Fasting Time in hours (mean (SD)) |  | 14.06 (1.89) | 14.16 (1.91) |
| Daily caffeine intake (mean (SD)) |  | 100.86 (116.68) | 106.60 (120.97) |
| WHIIRS (mean (SD)) |  | 7.13 (5.36) | 7.72 (5.53) |
| Suspected Insomnia (%) | Yes | 1,411 (33.2) | 892 (37.1) |
| Excessive Daytime Sleepiness (ESS≥11) | Yes | 606 (14.7) | 353 (15.8) |
| Average Time in Bed in hours (mean (SD)) |  | 7.93 (1.45) | 7.87 (1.49) |
| Mild-to-severe OSA (%) | Yes | 1,088 (27.3) | 751 (36.5) |
| Shift Work (%) | Yes | 762 (20.1) | 328 (16.0) |
| Sleep Medication Usage | Yes | 539 (12.4) | 372 (18.1) |
| Suspected Depression (CESD10≥10) | Yes | 1,168 (25.7) | 718 (31.4) |
| Elevated Anxiety (STAI10≥20) | Yes | 1,140 (27.0) | 661 (27.4) |
| Mild Cognitive Impairment | Yes | 170 (11.5) | 185 (11.3) |

The numbers are unweighted, while means and percentages are weighted to be representative of the HCHS/SOL target population. Abbreviations: N=Number; %=Percentage; SD= Standard Deviation; AHEI = Alternate Healthy Eating Index; WHIIRS = Women's Health Initiative Insomnia Rating Scale; ESS = Epworth Sleepiness Scale; OSA = Obstructive Sleep Apnea; CESD10 = Center for Epidemiologic Studies Depression Scale short form; STAI10 = State-Trait Anxiety Inventory short form

### Metabolite-wide association study (MWAS) for suspected insomnia and insomnia symptoms (WHIIRS)

Statistically significant associations from the discovery analysis in batch 1 and the corresponding results from replication analysis in batch 2 are presented in Table 2. Complete MWAS results for insomnia symptoms (WHIIRS) and suspected insomnia are available in Supplementary Tables 2 and 3, respectively. No associations with the dichotomized phenotype, suspected insomnia, passed the multiple testing threshold. However, nine metabolites were statistically significantly associated with WHIIRS at an FDR p-value of less than 0.05. In the replication analysis, only two metabolite associations were replicated: hydrocinnamate (FDR p = 0.05) and indolepropionate (FDR p = 0.002). Higher levels of these metabolites were associated with lower WHIIRS values (fewer insomnia symptoms). As the metabolites were rank-normalized, effect sizes are difficult to interpret. We visualize the estimated effect sizes and corresponding 95% confidence intervals in Figure 2 using the pooled batch-combined data, both in our primary analysis and with different covariate adjustments (Models 1-4), and excluding medication users in the sensitivity analysis.

**Table 2:** Statistically significant metabolite associations with WHIIRS in primary analysis.

| Metabolite Name | Batch 1 Discovery Analysis (n=3,932) |  |  |  |  | Batch 2 Replication Analysis (n=2,175) |  |  |  |  |
| --- | --- | --- | --- | --- | --- | --- | --- | --- | --- | --- |
|  | Beta | 95% CI |  | P-value | FDR-P | Beta | 95% CI |  | P-value | FDR-P |
|  |  | LL | UL |  |  |  | LL | UL |  |  |
| indolepropionate | -0.015 | -0.023 | -0.007 | 3E-04 | 0.03 | -0.018 | -0.028 | -0.008 | 2E-04 | 0.002 |
| 3-phenylpropionate (hydrocinnamate) | -0.016 | -0.024 | -0.009 | 4E-05 | 0.01 | -0.012 | -0.023 | -0.002 | 0.01 | 0.05 |
| hydroxy-CMPF | -0.017 | -0.025 | -0.009 | 7E-05 | 0.01 | -0.008 | -0.017 | 0.002 | 0.06 | 0.18 |
| 1-palmitoyl-2-palmitoleoyl-GPC (16:0/16:1) | 0.014 | 0.006 | 0.022 | 4E-04 | 0.03 | 0.006 | -0.003 | 0.015 | 0.10 | 0.22 |
| 1-palmitoleoyl-GPC (16:1) | 0.015 | 0.007 | 0.024 | 3E-04 | 0.03 | 0.005 | -0.005 | 0.015 | 0.16 | 0.30 |
| 3-bromo-5-chloro-2,6-dihydroxybenzoic acid | -0.015 | -0.022 | -0.008 | 4E-05 | 0.01 | -0.005 | -0.017 | 0.007 | 0.21 | 0.31 |
| 3-carboxy-4-methyl-5-propyl-2-furanpropanoate (CMPF) | -0.016 | -0.024 | -0.008 | 4E-05 | 0.01 | -0.002 | -0.011 | 0.008 | 0.35 | 0.35 |
| 3,5-dichloro-2,6-dihydroxybenzoic acid | -0.015 | -0.022 | -0.008 | 7E-05 | 0.01 | -0.002 | -0.014 | 0.009 | 0.34 | 0.35 |
| 1-palmitoyl-2-oleoyl-GPC (16:0/18:1) | 0.013 | 0.006 | 0.021 | 4E-04 | 0.03 | 0.002 | -0.008 | 0.012 | 0.34 | 0.35 |

The table presents association analysis results for metabolites with an FDR-corrected p-value (FDR-P) of less than 0.05 in the batch 1 discovery analysis and their subsequent replication analysis in batch 2. A metabolite association is considered replicated with the FDR-P less than 0.05 in batch 2. Beta is the regression (beta) estimate of WHIIRS association from the survey regression of the rank-normalized metabolite level over WHIIRS, adjusted for Model 1 covariates: age, sex, study center, and self-reported Hispanic/Latino background. Abbreviations: CI, Confidence Interval; LL, Lower Limit; UL, Upper Limit; FDR-P, false discovery rate P-value; WHIIRS, Women's Health Initiative Insomnia Rating Scale.

**Figure 2.**
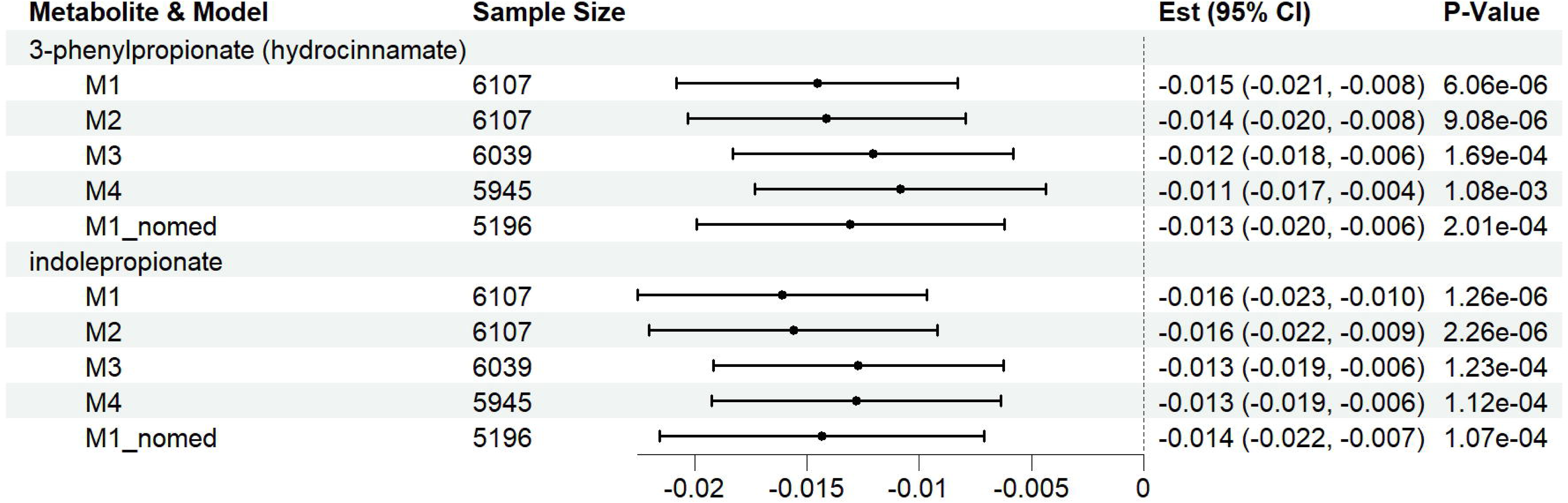
Estimated associations of the metabolites with WHIIRS-replicated associations with WHIIRS in batch combined analysis across regression models. The figure visualizes the estimated associations of hydrocinnamate and indolepropionate with WHIIRS in four analysis models. Model 1 (M1) was adjusted for age, sex, study center, and Hispanic/Latino background. Model 2 (M2) adjusted for M1 covariates and diabetes and hypertension status. Model 3 adjusted for M2 covariates and BMI, alcohol use, smoking status, physical activity, and diet (AHEI 2010). Model 4 adjusted for M3 and additionally for daily caffeine intake and fasting time before the blood sampling. The M1_nomed model is the same as Model 1, excluding individuals who reported using sleep medication. “Est” is the estimated effect size from the regression of WHIIRS as an outcome over the metabolite and covariates, and CI is the 95% confidence interval. P-value is the raw p-value from the 1-degree-of-freedom Wald test. Abbreviations: AHEI, Alternative Health Eating Index; BMI, Body Mass Index; CI, Confidence Interval; FDR, False Discovery Rate; WHIIRS, Women’s Health Initiative Insomnia Rating Scale.

### Metabolite risk scores for insomnia symptoms (WHIIRS) and suspected insomnia

Using data from batch 1, we developed MRSs for insomnia symptoms and suspected insomnia. Supplementary Table 4 provides the selected metabolites and their estimated coefficients from LASSO for each MRS. Figure 3 and Supplementary Table 5 provide the associations in the validation dataset (batch 2) of the two MRSs - one including all individuals and one excluding those who reported sleep medication use - with suspected insomnia, insomnia symptoms (WHIIRS), and the subcomponent questions. The MRS for suspected insomnia has 61 when all individuals are considered and 77 metabolites when excluding sleep medication users. The insomnia symptoms (WHIIRS) MRS has 51 and 43 metabolites, respectively. Of the two metabolites associated with insomnia symptoms in the MWAS analysis, only indoleproprionate was included in the insomnia MRS using the total population of batch 1. For the WHIIRS MRS, both metabolites were included for both total population and when excluding sleep medication users. In the replication study (batch 2), the odds ratio (OR) for suspected insomnia per one standard deviation (SD) increase was 1.28 (95% CI [1.11, 1.49]) among all studied participants, and 1.23 (95% CI [1.07, 1.42]) when excluding sleep medication users. For insomnia symptoms, the association was similarly replicated in batch 2, and the effect size was weaker when excluding medication users from both the MRS construction dataset and the association analysis in batch 2.

**Figure 3:**
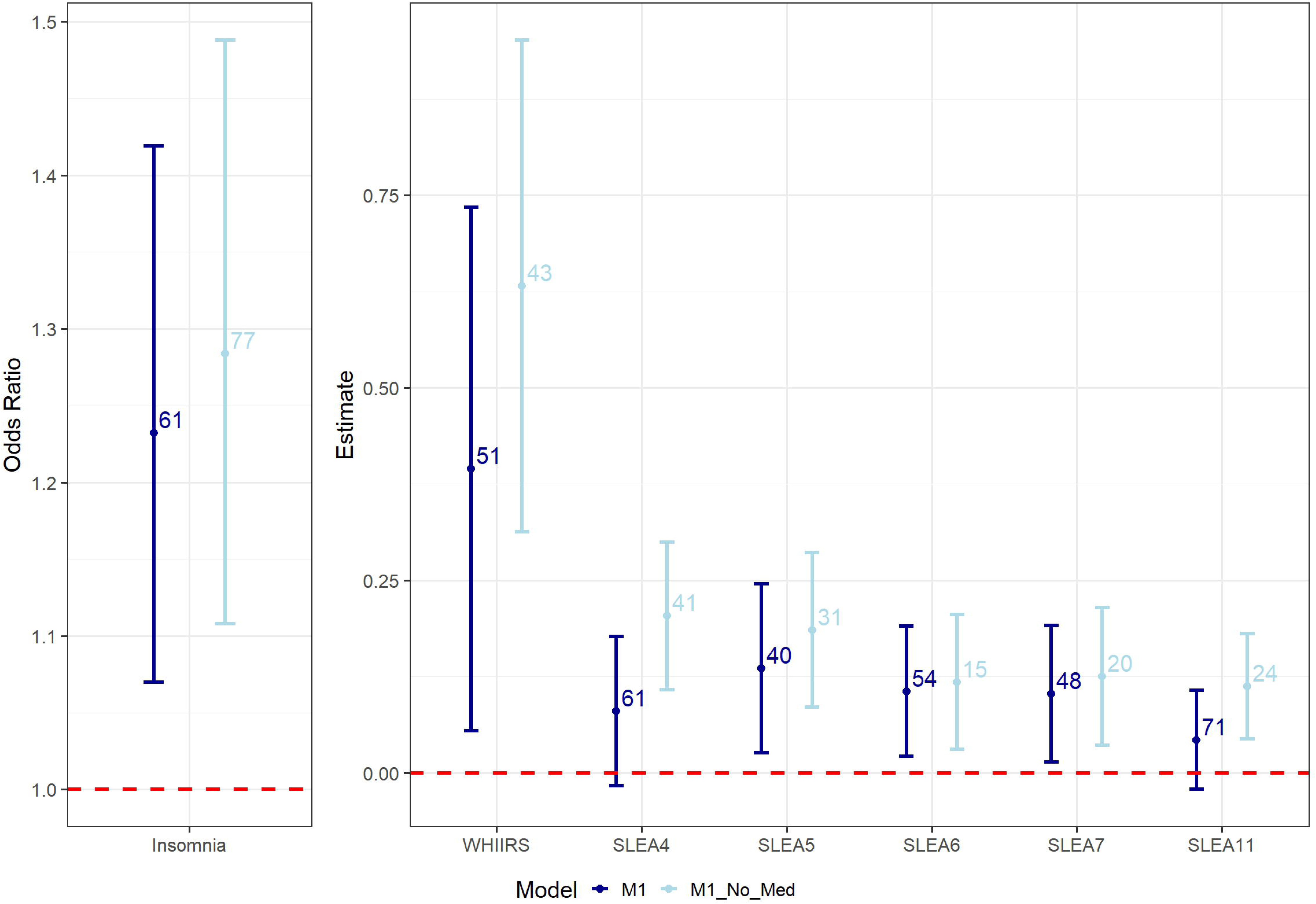
MRS associations with their insomnia phenotypes in batch 2 participants. The figure visualizes the associations of MRSs for insomnia symptom measures with their respective phenotypes. Two MRSs are provided for each phenotype: one based on all available individuals (M1) and another based on individuals excluding medication users (M1_No_Med). MRSs were developed in the batch 1 dataset for each insomnia-related phenotype. Associations and 95% confidence intervals are reported based on analysis in batch 2. The left panel corresponds to suspected insomnia (WHIIRS≥ 9) and provides the odds ratio from logistic regression. The right panel corresponds to continuous measures, where SLEA4-7 & SLEA11 are the subcomponents of the WHIIRS. SLEA4: Did you have trouble falling asleep? SLEA5: Did you wake up several times at night? SLEA6: Did you wake up earlier than you planned to? SLEA7: Did you have trouble getting back to sleep after waking up too early? SLEA11: Overall, your typical night’s sleep during the past 4 weeks was: (choices on a scale from 0 = very sound or restful to 4 = very restless). Abbreviations: MRS, metabolite risk scores; WHIIRS, Women’s Health Initiative Insomnia Rating Scale.

### Subcomponent analysis

Supplementary Table 6 presents the results from an association analysis of the two replicated WHIIRS-associated metabolites with the component questions of WHIIRS. We adjusted for age, sex, study center, and Hispanic/Latino background in the pooled batch-combined dataset. All sleep characteristics were associated with the two metabolites (indolepropionate and 3-phenylpropionate (hydrocinnamate)), with the same negative direction of association for all subcomponent questions. For indolepropionate and hydrocinnamate, the strongest estimated effect was observed in response to the question about sleep restfulness, indicating a 0.09-point decrease in WHIIRS for each one-point increase on the Likert scale, indicating more restless sleep. In addition, we also created MRS for the individual subcomponents of the WHIIRS (see Supplementary Table 4 for the coefficients and Supplemental Table 5 for the validation in batch 2). For these subcomponents, the majority of the MRS associations were statistically significant (p-value<0.05) in the replication dataset (batch 2). However, associations for two questions (trouble falling asleep and restlessness of their sleep) substantially weakened among the subgroup when excluding individuals who reported sleep medication. The number of metabolites in the MRS was lower among those who did not report sleep mediation (41 vs. 61 for trouble falling asleep, and 24 vs. 71 for restlessness). As sleep medication is highly correlated with reporting sleep onset problems and restfulness of sleep, this may be a reason why the results were so severely attenuated. Among individuals in batch 2 who reported severe trouble falling asleep (5+ days per week), 41.4% reported using sleep medication; compared to those who reported no difficulty falling asleep (no days per week), only 6.4 % reported sleep medication. Similarly, among individuals in batch 2 who reported very restless sleep, 37.1% reported using sleep medication; compared to those who reported no trouble falling asleep (no days per week), only 5.0 % reported sleep medication. The patterns in the individuals in batch 1 are similar.

### Cross-phenotype analyses

We report the results of the association between several other phenotypes and the two identified metabolites (hydrocinnamate and indolepropionate) that were associated with and replicated in insomnia symptoms, as well as the two (WHIIRS and suspected insomnia) MRS, in Table 3. These phenotypes include sleep-related, psychological, and cognitive phenotypes and were adjusted for age, sex, study center, and Hispanic/Latino background in a batch-combined analysis. Supplementary Table 7 provides the corresponding association with the various phenotypes for both metabolites and the two MRS, along with the association analyses that exclude medication users. Supplementary Table 8 only displays the results from the validation dataset (batch 2 participants). The effect size estimates for these subgroup analyses are very similar to those including the total population. The P-values were generally larger due to a more limited sample size.

**Table 3:**
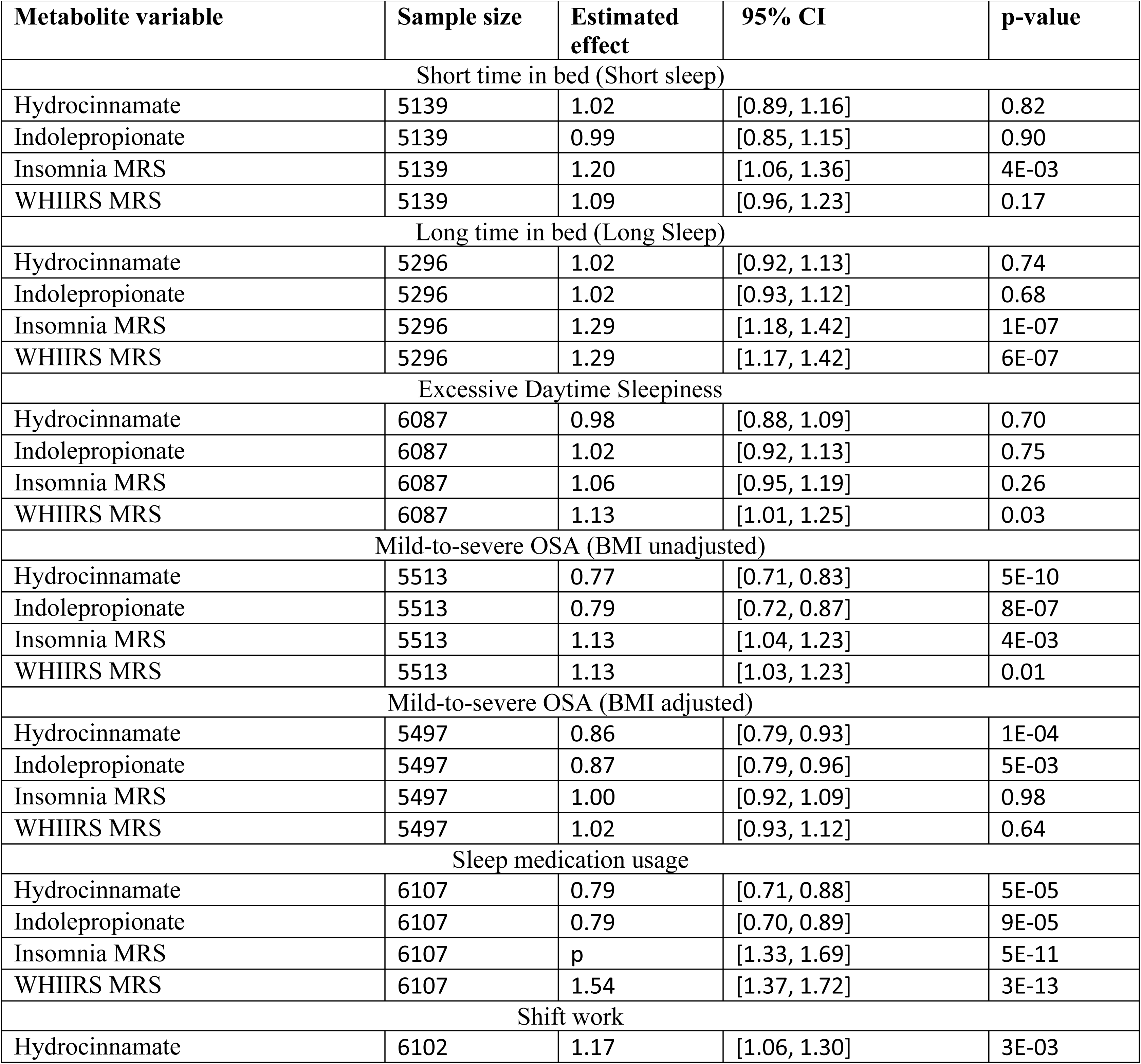
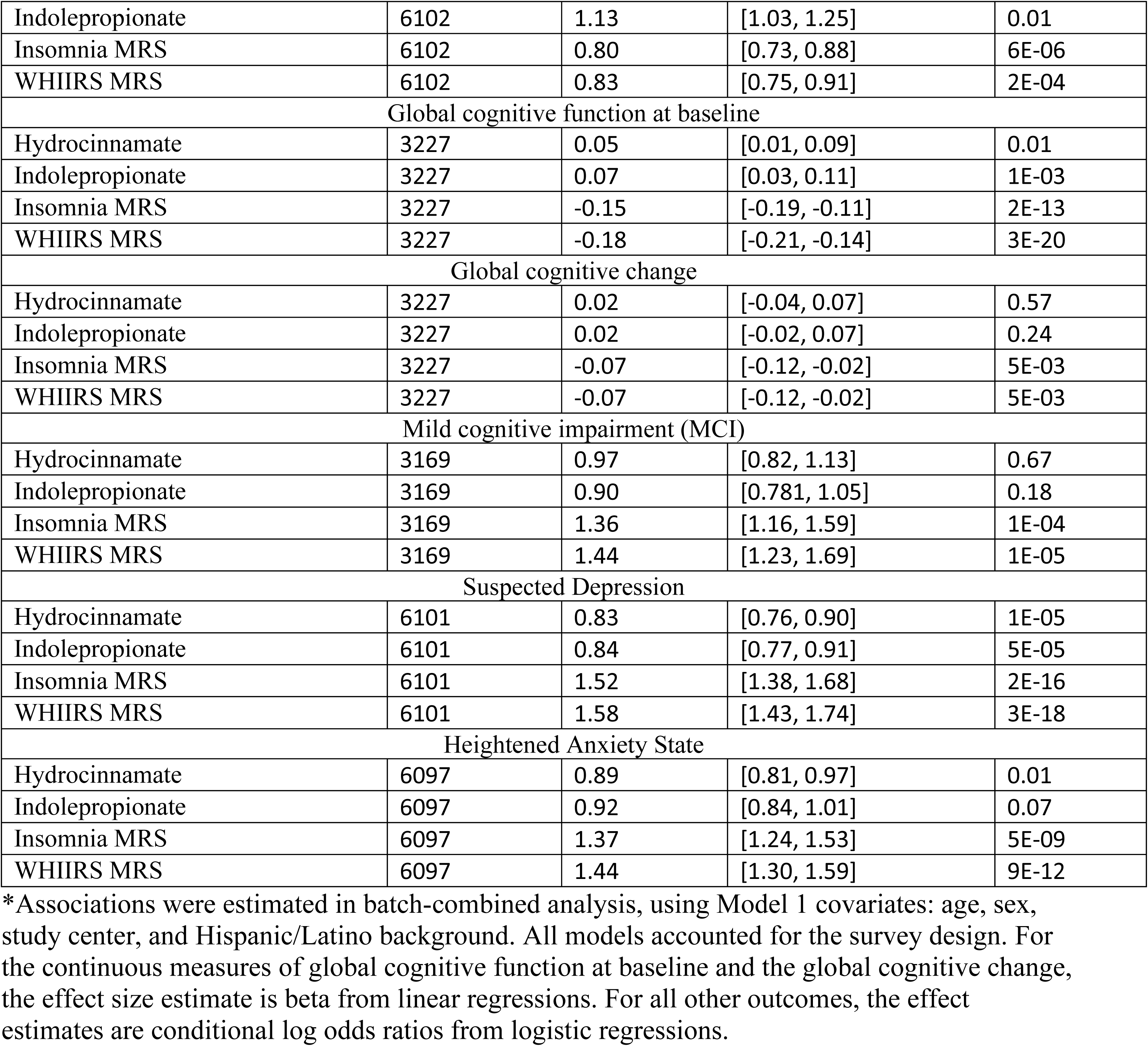
Estimated associations of WHIIRS-associated metabolite measures with other sleep and cognitive phenotypes.

As anticipated, given the inverse associations of hydrocinnamate and indolepropionate with insomnia symptoms, higher levels of these metabolites were also associated with a reduced risk of mild-to-severe obstructive sleep apnea (OSA), a lower likelihood of sleep medication use, and an increased likelihood of shift work. Similarly, the directions for the MRS were as expected, given that higher MRS assumes higher insomnia symptoms; the results for the MRS were positively associated with various sleep phenotypes. Higher insomnia and WHIIRS MRS values, indicating a higher risk for suspected insomnia or more insomnia symptoms, were associated with an increased likelihood of long sleep duration, use of sleep medication, a lower likelihood of shift work, and a higher likelihood of having mild-to-severe OSA. However, no association was observed in the BMI-adjusted analysis. Additionally, insomnia MRS was associated with a higher risk of short sleep duration, and the WHIIRS MRS showed a positive association with excessive daytime sleepiness.

In addition to associations with sleep-related phenotypes, where higher levels of hydrocinnamate and indolepropionate were linked to fewer insomnia symptoms, we also observed higher global cognitive function at baseline, and a lower risk for suspected depression or heightened anxiety status. At the follow-up visit, no associations were detected with cognitive change or mild cognitive impairment. Both the suspected insomnia and WHIIRS MRS were associated with an increased risk of suspected depression, heightened anxiety status, as well as lower cognitive function at baseline, increased cognitive decline from baseline, and the risk for mild cognitive impairment at the follow-up visit.

### Sensitivity analyses

For the MWAS analysis, we analyzed the two identified metabolites (hydrocinnamate and indolepropionate) with different covariate adjustments (Models 1-4), and when excluding medication users. These results are from the batch-combined analysis, and the estimated associations are relatively consistent across analyses (see Figure 2). In addition, analysis excluding individuals with only partial WHIIRS information indicated very similar results (Supplementary Table 9). Finally, we conducted batch-combined, sex-combined, and sex-stratified MWAS analyses. The results are reported in Supplementary Tables 10 (WHIIRS) and 11 (suspected insomnia). For suspected insomnia and WHIIRS, we used an FDR adjusted P-value for each sex separately, and calculated a Q-statistic to test for heterogeneity. No metabolite was statistically significant for suspected insomnia in the total or female-only population. Among males, higher indolepropionate was associated with a decreased risk for suspected insomnia. Among the pooled total dataset, 26 metabolites were statistically significant with insomnia symptoms (WHIIRS). For males only, one of the 26 metabolites was statistically significant (indolepropionate), and one additional metabolite that was not statistically significant in the total population was identified to be associated with lower insomnia symptoms (X-11880). Only one of the 26 metabolites among females was statistically significant (hydroxy-CMPF). No metabolites were statistically significantly different (Q-test: FDR-P<0.05) between the two sexes. For the 26 metabolites identified in the total population, the direction of the association was in the same direction for both males and females, even though the effect estimates were different. Many of these metabolites are associated with diet and the gut microbiome. For example, according to previous reports, hydroxy-CMPF and carotene diol are metabolites strongly related to diet.[55–57] Additionally, glycoursodeoxycholic acid, glycochenodeoxycholate, and glycocholate are associated with gut microbiome and fat intake [58] and have previously been proposed to reduce the risk of type 2 diabetes in mice.[59]

## Discussion

We performed an association analysis of blood metabolites and insomnia symptoms in a large cohort of Hispanic/Latino adults in the U.S. Using a discovery-replication approach in our primary MWAS analysis, we leveraged separate subsets of the HCHS/SOL with metabolites from different assay batches. We identified two replicated metabolite associations with insomnia symptoms based on WHIIRS: hydrocinnamate and indolepropionate. We then developed MRS for suspected insomnia and insomnia symptoms using batch 1 and validated them in the second batch. The WHIIRS and insomnia MRS, as well as the two metabolites identified in the MWAS, were associated with various other sleep phenotypes, suspected depression and heightened anxiety, and several cognitive outcomes, including lower cognitive function at baseline cross-sectionally and mild cognitive impairment on average seven years later, suggesting that an insomnia-specific metabolic profile may partly capture insomnia’s link to cognitive aging.

Hydrocinnamate, also known as 3-phenylpropanoic acid, is a xenobiotic metabolite produced in the gut microbiome through the metabolism of dietary polyphenols, primarily sourced from fruit. [60,61] The gut microbiome reduces hydroxycinnamatases - well-known antioxidants - [60] into hydrocinnamate (3-phenylpropionic acid), [61] which plays a role in gut health and muscle function/growth. [62,63] In our analysis, higher hydrocinnamate levels were associated with fewer insomnia symptoms, a lower likelihood of OSA, and reduced sleep medication use. These associations may reflect the established link between healthy sleep and a diet rich in polyphenols.[64] This is consistent with previous research showing that an anti-inflammatory diet, characterized by higher antioxidant intake including hydrocinnamate, is associated with a reduced risk of OSA.[65] Interestingly, higher hydrocinnamate levels were associated with a greater likelihood of being a shift worker. While this initially appears paradoxical, the elevated hydrocinnamate levels observed among shift workers may reflect a behavioral adaptation to the demands of shift work. Individuals who are more resilient or better adapted to irregular schedules may develop specific dietary patterns, such as increased consumption of coffee (which is high in caffeic acid and metabolized to hydrocinnamate). Shift workers typically consume more coffee,[66] and hypothetically, they would experience fewer adverse effects of coffee on insomnia symptoms. This could result in a higher overall hydrocinnamate intake, despite their generally lower consumption of vegetables and fruits.[67]

Indolepropionate, also known as indole-3 propionate, is an amino acid from the tryptophan metabolism pathway. It is produced in the gut by interacting with tryptophan and the gut microbiome.[68] While previous studies have identified associations between tryptophan concentrations and lower insomnia symptoms,[65] our analysis found no association between tryptophan levels themselves and suspected insomnia or insomnia symptoms. Higher indolepropionate concentration is indicative of greater microbiome diversity.[69] It is associated with increased fruit intake and reduced egg and red meat consumption.[70] In addition, several health conditions have been associated with indolepropionate, including better muscle strength, improved insulin sensitivity, lower inflammation, and a lower incidence of diabetes and kidney diseases.[70–75] In rodent studies, dietary supplementation has been shown to enhance metabolic health, lower inflammation, and exhibit neuroregenerative properties.[76–79] A trial (NCT06674018) was initiated in February 2025 to review the safety and anti-inflammatory response to daily supplementation of indole propionate in healthy individuals. Results are pending.

In our data, higher levels of indolepropionate were linked to fewer insomnia symptoms, reduced use of sleep medications, a lower risk of OSA, lower depression symptoms, and better global cognitive function at baseline. These associations may reflect indolepropionate’s broader metabolic and anti-inflammatory effects. Like hydrocinnamate, higher concentrations of indolepropionate are associated with an increased risk of shift work. This can be explained by the notion that shift workers may be somewhat protected against insomnia, as individuals more prone to insomnia symptoms might be more likely to choose day shifts over rotating or night shifts. Finally, the overlap of these two metabolites, both hydrocinnamate and indolepropionate, with both insomnia and OSA may help explain the co-occurrence of these disorders, known as COMISA – co-morbid insomnia and sleep apnea.[80]

MRS are generally more powerful than single metabolites in summarizing the metabolic profile of a phenotype, as they utilize information from multiple metabolites simultaneously. While more challenging to interpret, MRS can be a useful overall biomarker. For example, we previously demonstrated that MRS for sleep disordered breathing phenotypes has stronger associations with incident hypertension and diabetes than measured physiological traits.[33] In this study, we explored the associations between suspected insomnia and insomnia symptoms (WHIIRS) MRS with additional sleep-related, psychological, and cognitive phenotypes. We found several associations, with consistent directions across the MRS. Specifically, higher MRS values (more insomnia symptoms) were associated with a higher likelihood of long sleep duration, an increased risk of OSA (but only in BMI-unadjusted analysis), more frequent use of sleep medication, a lower likelihood of being a shift worker, higher anxiety and suspected depression, lower global cognitive function at baseline, a negative change in global cognitive function, and a greater likelihood of having MCI approximately seven years after the baseline exam. Since MRS captures more comprehensive metabolic data than single metabolites, most of these findings with comorbid measures were statistically stronger than the two individual metabolites. There are inconsistencies between the results of single-metabolite associations and MRS associations. For example, hydrocinnamate and indolepropionate are associated with OSA, but the MRS revealed a weaker association that becomes null when adjusting for BMI. One possible explanation is that hydrocinnamate and indolepropionate may specifically target COMISA, whereas the MRS might indicate a broader aspect of insomnia. An unexpected finding from the study was the relationship between insomnia and insomnia symptoms MRS and long sleep duration. However, only the insomnia MRS was associated with short sleep. This may be related to how we define sleep duration based on bed and wake times. Individuals with insomnia often have poor sleep quality but might spend a longer time in bed to compensate. In addition, these findings could suggest that the biological underpinnings of insomnia, as captured by MRS, may relate differently to insomnia with varying sleep durations, potentially reflecting distinct physiological or behavioral pathways.

We have previously studied the associations of insomnia with cognitive outcomes in the same HCHS/SOL cohort. For example, in a causal association analysis, we found that individuals with suspected insomnia at the baseline HCHS/SOL exam were more likely to develop MCI at the SOL-INCA visit (on average, 7 years after baseline) when using methods designed to “equalize” the distribution of confounders between the exposed and unexposed groups.[81] In another study within HCHS/SOL, we identified that a higher genetic liability to insomnia, as assessed through a polygenic risk score (PRS), was associated with lower cognitive function at baseline and a higher likelihood of having MCI at the follow-up visit.[49] Compared to the (conditional) odds ratio for MCI per 1 SD increase in the PRS (OR = 1.20, 95% CI [1.06, 1.35], p-value = 0.005), the estimated effect size for the insomnia MRS was larger (OR=1.36, 95% CI [1.16, 1.59], p-value=0.0001). This slightly stronger association for the MRS may reflect its sensitivity to both genetic and environmental factors. In contrast, the PRS captures only inherited genetic predisposition. These findings indicate that the two scores capture complementary aspects of biological vulnerability—genetics representing fixed susceptibility, and metabolomics potentially marking more dynamic, modifiable risk scores.

In other work within the HCHS/SOL and Atherosclerosis Risk in Communities (ARIC) cohorts, we conducted a metabolomics analysis on global cognitive function.[82] We identified several dietary metabolites, primarily sugars, associated with cognitive performance. Some of these metabolites also had a suggestive association in our study. For instance, beta-cryptoxanthin has been associated with higher cognitive function and fewer insomnia symptoms (Supplementary Table 11), and some of the cognitive function-associated metabolites are included in the MRS developed in this study (Supplementary Table 5). As genetic and metabolomics data and methodologies continue to evolve and become more accessible, both the insomnia PRS and MRS are likely to improve with better selection of genetic variants and metabolites. This could lead to the development of more accurate association models and potentially even prediction models that can be calibrated across various populations. As models and pathway analysis continue to enhance, the goal of identifying specific pathways for targeting interventions to alleviate insomnia symptoms and improve cognitive function becomes more achievable.

This study focused on the understudied population of Hispanic/Latino adults in the U.S., and research on sleep health within this population is crucial for several reasons. Hispanic/Latino individuals represent the largest racial/ethnic minority population in the U.S., and are projected to make up more than 26% of the population by 2060.[71] As a result, the incidence of Alzheimer’s Disease (AD) and AD-related dementia is projected to increase more among this population than in any other U.S. ethnic or minority group.[72] Previous analyses in the Health and Retirement Study revealed that Hispanic adults in the U.S. have a steeper increase in insomnia symptoms over time compared to non-Hispanic White adults.[4]. However, since our analysis concentrated on this population, these findings should be interpreted cautiously when considering other ethnic or racial groups, as further validation in additional populations is needed.

In addition to involving this understudied population, other strengths of this analysis include a large, well-characterized sample with an extensive set of metabolites and a robust statistical methodology, featuring a discovery-replication design. This study does have certain limitations. For instance, we did not utilize the entire sample from the metabolomics data within HCHS/SOL in the primary analysis because we preferred using a discovery-validation dataset and could not replicate the results in another dataset. The insomnia phenotype is not a clinical measure but based on a self-reported scale. It is important to note that while the WHIIRS includes questions about trouble with sleep onset, maintenance, early wakings, and morning restfulness, it does not explicitly assess the daytime consequences of these sleep difficulties. Nevertheless, WHIIRS is a well-validated measure with good reliability and validity that has been employed in numerous research studies across various populations.[36,38] Therefore, although this is not a clinical diagnosis, this questionnaire is highly effective for assessing insomnia symptoms in a large research setting. Finally, because of the cross-sectional and observational nature of the data, we cannot draw any conclusions about the direction and causality of these associations.

In summary, we investigated the associations between metabolites and insomnia phenotypes, other sleep phenotypes, anxiety, depression symptoms, and cross-sectional and prospective global cognition measures in a comprehensive study involving Hispanic/Latino individuals. Future research should further assess the generalizability of these associations to other populations and utilize metabolomics to identify potential causal pathways, such as metabolites downstream of diet, like hydrocinnamate and indolepropionate, in relation to insomnia. Supplementation could be investigated as a potential therapeutic strategy if these metabolites are confirmed to play a causal role and are safe.

## Data sharing agreement

HCHS/SOL metabolomics and phenotype data are available via data use agreement with the HCHS/SOL Data Coordinating Center (DCC) at the University of North Carolina at Chapel Hill, see collaborators’ website: https://sites.cscc.unc.edu/hchs/. The code used in this work is available in the GitHub repository at https://github.com/tamartsi/Insomniablites.

## Acknowledgements

The authors thank the staff and participants of HCHS/SOL for their essential contributions—investigators’ website - http://www.cscc.unc.edu/hchs/. Grants from various National Institutes of Health and the JLH Foundation supported the work. Specifically, the National Heart Lung and Blood Institute (NHLBI) grants R01HL161012 to T.S., R35HL135818 to S.R., National Institute on Aging (NIA) grant R01AG080598 to T.S., and NIA grant R01AG067568 to ARR; and NIA grant K01AG081542 to CK. Support for metabolomics data was graciously provided by the JLH Foundation (Houston, Texas) and by NHLBI grant R01HL141824. The Hispanic Community Health Study/Study of Latinos is a collaborative study supported by contracts from the National Heart, Lung, and Blood Institute (NHLBI) to the University of North Carolina (HHSN268201300001I / N01-HC-65233), University of Miami (HHSN268201300004I / N01- HC-65234), Albert Einstein College of Medicine (HHSN268201300002I / N01-HC-65235), University of Illinois at Chicago (HHSN268201300003I / N01- HC-65236 Northwestern Univ), and San Diego State University (HHSN268201300005I / N01-HC-65237). The following Institutes/Centers/Offices have contributed to the HCHS/SOL through a transfer of funds to the NHLBI: National Institute on Minority Health and Health Disparities, National Institute on Deafness and Other Communication Disorders, National Institute of Dental and Craniofacial Research, National Institute of Diabetes and Digestive and Kidney Diseases, National Institute of Neurological Disorders and Stroke, NIH Institution-Office of Dietary Supplements. In addition, the National Institute on Aging provided support for SOL-INCA (R01AG048642).

## Author contributions

TS conceptualized the study. JSC, Yu. Z, CK, and TS performed analyses. Yu. Z and CK prepared final tables and figures and finalized the code. CK, JSC, and TS drafted the manuscript. BY and QQ designed the metabolomics data collection. ARR, MD, PCZ, RK, HMG, CRI, and SR designed or contributed to HCHS/SOL data collection, including sleep and cognitive data. JSC, CK, Yu.Z, TH, JC, RA, BY, QQ, CA, ARR, MD, PCS, RK, HMG, CRI, SR, and TS all critically reviewed the manuscript. TS supervised the work.

## References

[1] Riemann D, Fee Benz |, Dressle RJ, Espie CA, Johann AF, Blanken TF, et al. Insomnia disorder: State of the science and challenges for the future. J Sleep Res 2022. 10.1111/jsr.13604.

[2] LeBlanc M, Beaulieu-Bonneau S, Mérette C, Savard J, Ivers H, Morin CM. Psychological and health-related quality of life factors associated with insomnia in a population-based sample. J Psychosom Res 2007;63:157–66. 10.1016/j.jpsychores.2007.03.004.

[3] Morin CM, Jarrin DC. Epidemiology of Insomnia: Prevalence, Course, Risk Factors, and Public Health Burden. Sleep Med Clin 2022;17. 10.1016/J.JSMC.2022.03.003.

[4] Kaufmann CN, Mojtabai R, Hock RS, Thorpe RJ, Canham SL, Chen LY, et al. Racial/Ethnic Differences in Insomnia Trajectories among U.S. Older Adults. American Journal of Geriatric Psychiatry 2016;24:575–84. 10.1016/j.jagp.2016.02.049.

[5] Andersen ML, Poyares D, Tufik S. Insomnia and cardiovascular outcomes. Sleep Science 2021;14:1–2. 10.5935/1984-0063.20200109.

[6] Yuan S, Mason AM, Burgess S, Larsson SC. Genetic liability to insomnia in relation to cardiovascular diseases: a Mendelian randomisation study. Eur J Epidemiol 2021;36:393– 400. 10.1007/s10654-021-00737-5.

[7] Wardle-Pinkston S, Slavish DC, Taylor DJ. Insomnia and cognitive performance: A systematic review and meta-analysis. Sleep Med Rev 2019;48. 10.1016/j.smrv.2019.07.008.

[8] Huang J, Spira AP, Perrin NA, Ellis A, Hsu EC, Kaufmann CN, et al. Latent classes of sleep problems and subjective cognitive decline among middle-aged and older adults in the United States. Arch Gerontol Geriatr 2025;129. 10.1016/j.archger.2024.105657.

[9] de Almondes KM, Costa MV, Malloy-Diniz LF, Diniz BS. Insomnia and risk of dementia in older adults: Systematic review and meta-analysis. J Psychiatr Res 2016;77:109–15. 10.1016/j.jpsychires.2016.02.021.

[10] Sorlie PD, Allison MA, Avilés-Santa ML, Cai J, Daviglus ML, Howard AG, et al. Prevalence of Hypertension, Awareness, Treatment, and Control in the Hispanic Community Health Study/Study of Latinos. Am J Hypertens 2014;27:793–800. 10.1093/AJH/HPU003.

[11] Zissimopoulos JM, Tysinger BC, St Clair PA, Crimmins EM. The Impact of Changes in Population Health and Mortality on Future Prevalence of Alzheimer’s Disease and Other Dementias in the United States. J Gerontol B Psychol Sci Soc Sci 2018;73:S38. 10.1093/GERONB/GBX147.

[12] Daviglus ML, Talavera GA, Avilés-Santa ML, Allison M, Cai J, Criqui MH, et al. Prevalence of Major Cardiovascular Risk Factors and Cardiovascular Diseases Among Hispanic/Latino Individuals of Diverse Backgrounds in the United States. JAMA 2012;308:1775–84. 10.1001/JAMA.2012.14517.

[13] Issaq HJ, Van QN, Waybright TJ, Muschik GM, Veenstra TD. Analytical and statistical approaches to metabolomics research. J Sep Sci 2009;32:2183–99. 10.1002/jssc.200900152.

[14] Gertsman I, Barshop BA. Promises and pitfalls of untargeted metabolomics. J Inherit Metab Dis 2018;41:355–66. 10.1007/s10545-017-0130-7.

[15] Gonzalez-Covarrubias V, Martínez-Martínez E, Bosque-Plata L Del. The Potential of Metabolomics in Biomedical Applications. Metabolites 2022;12. 10.3390/metabo12020194.

[16] Diallo I, Pak VM. Metabolomics, sleepiness, and sleep duration in sleep apnea. Sleep and Breathing 2020;24:1327–32.

[17] Xu H, Zheng X, Jia W, Yin S. Chromatography/mass spectrometry-based biomarkers in the field of obstructive sleep apnea. Medicine (United States) 2015;94:1–9. 10.1097/MD.0000000000001541.

[18] Conte L, Greco M, Toraldo DM, Arigliani M, Maffia M, De Benedetto M. A review of the “omics” for management of patients with obstructive sleep apnoea. Acta Otorhinolaryngologica Italica 2020;40:164–72. 10.14639/0392-100X-N0409.

[19] Humer E, Pieh C, Brandmayr G. Metabolomics in Sleep, Insomnia and Sleep Apnea. Int J Mol Sci 2020;21:1–17. 10.3390/IJMS21197244.

[20] Zhang Y, Ngo D, Yu B, Shah NA, Chen H, Ramos AR, et al. Development and validation of a metabolite index for obstructive sleep apnea across race/ethnicities. Sci Rep 2022;12. 10.1038/s41598-022-26321-9.

[21] Zhang Y, Spitzer BW, Zhang Y, Wallace DA, Yu B, Qi Q, et al. Untargeted metabolome atlas for sleep-related phenotypes in the Hispanic community health study/study of Latinos. EBioMedicine 2025;111:105507. 10.1016/j.ebiom.2024.105507.

[22] Xiao Q, Derkach A, Moore S, Zheng W, Shu X, Gu F, et al. Habitual Sleep and human plasma metabolomics. Metabolomics 2017;13:63. 10.1007/S11306-017-1205-Z.

[23] Honma A, Revell VL, Gunn PJ, Davies SK, Middleton B, Raynaud FI, et al. Effect of acute total sleep deprivation on plasma melatonin, cortisol and metabolite rhythms in females. European Journal of Neuroscience 2020;51:366–78. 10.1111/ejn.14411.

[24] Gordon-Dseagu VLZ, Derkach A, Xiao Q, Williams I, Sampson J, Stolzenberg-Solomon RZ. The association of sleep with metabolic pathways and metabolites: evidence from the Dietary Approaches to Stop Hypertension (DASH)—sodium feeding study. Metabolomics 2019 15:4 2019;15:1–14. 10.1007/S11306-019-1472-Y.

[25] Depner CCM, Cogswell DDT, Bisesi PPJ, Markwald RRR, Cruickshank-Quinn C, Quinn K, et al. Developing preliminary blood metabolomics-based biomarkers of insufficient sleep in humans. Sleep 2020;43:1–13. 10.1093/SLEEP/ZSZ321.

[26] Bhat A, Pires AS, Tan V, Babu Chidambaram S, Guillemin GJ. Effects of Sleep Deprivation on the Tryptophan Metabolism. International Journal of Tryptophan Research 2020;13:1178646920970902. 10.1177/1178646920970902.

[27] Topriceanu CC, Tillin T, Chaturvedi N, Joshi R, Garfield V. The association between plasma metabolites and sleep quality in the Southall and Brent Revisited (SABRE) Study: A cross-sectional analysis. J Sleep Res 2021;30:e13245. 10.1111/JSR.13245.

[28] Huang T, Zeleznik OA, Poole EM, Clish CB, Deik AA, Scott JM, et al. Habitual sleep quality, plasma metabolites and risk of coronary heart disease in post-menopausal women. Int J Epidemiol 2019;48:1262–74. 10.1093/ije/dyy234.

[29] Gehrman P, Sengupta A, Harders E, Ubeydullah E, Pack AIA, Weljie A. Altered diurnal states in insomnia reflect peripheral hyperarousal and metabolic desynchrony: A preliminary study. Sleep 2018;41:1–12. 10.1093/sleep/zsy043.

[30] Tanaka A, Sanada K, Miyaho K, Tachibana T, Kurokawa S, Ishii C, et al. The relationship between sleep, gut microbiota, and metabolome in patients with depression and anxiety: A secondary analysis of the observational study. PLoS One 2023;18. 10.1371/journal.pone.0296047.

[31] Bos MM, Goulding NJ, Lee MA, Hofman A, Bot M, Pool R, et al. Investigating the relationships between unfavourable habitual sleep and metabolomic traits: evidence from multi-cohort multivariable regression and Mendelian randomization analyses. BMC Med 2021;19. 10.1186/s12916-021-01939-0.

[32] Buergel T, Steinfeldt J, Ruyoga G, Pietzner M, Bizzarri D, Vojinovic D, et al. Metabolomic profiles predict individual multidisease outcomes. Nat Med 2022;28:2309– 20. 10.1038/s41591-022-01980-3.

[33] Zhang Y, Yu B, Qi Q, Azarbarzin A, Chen H, Shah NA, et al. Metabolomic profiles of sleep-disordered breathing are associated with hypertension and diabetes mellitus development. Nat Commun 2024;15. 10.1038/s41467-024-46019-y.

[34] Fritz J, Huang T, Depner CM, Zeleznik OA, Cespedes Feliciano EM, Li W, et al. Sleep duration, plasma metabolites, and obesity and diabetes: a metabolome-wide association study in US women. Sleep 2023;46:1–12. 10.1093/sleep/zsac226.

[35] Sorlie PD, Avilés-Santa LM, Wassertheil-Smoller S, Kaplan RC, Daviglus ML, Giachello AL, et al. Design and implementation of the Hispanic Community Health Study/Study of Latinos. Ann Epidemiol 2010;20:629–41. 10.1016/J.ANNEPIDEM.2010.03.015.

[36] Levine DW, Dailey ME, Rockhill B, Tipping D, Naughton MJ, Shumaker SA. Validation of the Women’s Health Initiative Insomnia Rating Scale in a multicenter controlled clinical trial. Psychosom Med 2005;67:98–104. 10.1097/01.PSY.0000151743.58067.F0.

[37] Levine DW, Kaplan RM, Kripke DF, Bowen DJ, Naughton MJ, Shumaker SA. Factor structure and measurement invariance of the Women’s Health Initiative Insomnia Rating Scale. Psychol Assess 2003;15:123–36. 10.1037/1040-3590.15.2.123.

[38] Levine DW, Kripke DF, Kaplan RM, Lewis MA, Naughton MJ, Bowen DJ, et al. Reliability and validity of the Women’s Health Initiative Insomnia Rating Scale 2003;15:137–48. 10.1037/1040-3590.15.2.137.

[39] Feofanova E V., Chen H, Dai Y, Jia P, Grove ML, Morrison AC, et al. A Genome-wide Association Study Discovers 46 Loci of the Human Metabolome in the Hispanic Community Health Study/Study of Latinos. The American Journal of Human Genetics 2020;107:849–63. 10.1016/J.AJHG.2020.09.003.

[40] Gonzalez S, Strizich G, Isasi CR, Hua S, Comas B, Sofer T, et al. Consent for Use of Genetic Data among US Hispanics/Latinos: Results from the Hispanic Community Health Study/ Study of Latinos. Ethn Dis 2021;31:547–58. 10.18865/ed.31.4.547.

[41] Qureshi WT, Leigh JA, Swett K, Dharod A, Allison MA, Cai J, et al. Comparison of echocardiographic measures in a hispanic/latino population with the 2005 and 2015 American society of echocardiography reference limits (The Echocardiographic Study of Latinos). Circ Cardiovasc Imaging 2016;9. 10.1161/CIRCIMAGING.115.003597.

[42] Redline S, Sotres-Alvarez D, Loredo J, Hall M, Patel SR, Ramos A, et al. Sleep-disordered breathing in Hispanic/Latino individuals of diverse backgrounds: The Hispanic Community Health Study/Study of Latinos. Am J Respir Crit Care Med 2014;189:335–44. 10.1164/rccm.201309-1735OC.

[43] Spielberger C. State-Trait Anxiety Inventory. The corsini encyclopedia of psychology, Hoboken, NJ, USA: John Wiley & Sons, Inc; 2010.

[44] Miloyan B, Eaton W. The Center for Epidemiologic Studies Depression (CES-D). Encyclopedia of gerontology and population aging, Springer International Publishing; 2019.

[45] Perreira KM, Gotman N, Isasi CR, Arguelles W, Castañeda SF, Daviglus ML, et al. Mental Health and Exposure to the United States: Key Correlates from the Hispanic Community Health Study of Latinos. J Nerv Ment Dis 2015;203:670. 10.1097/NMD.0000000000000350.

[46] González HM, Tarraf W, Fornage M, González KA, Chai A, Youngblood M, et al. A research framework for cognitive aging and Alzheimer’s disease among diverse US Latinos: Design and implementation of the Hispanic Community Health Study/Study of Latinos—Investigation of Neurocognitive Aging (SOL-INCA). Alzheimer’s and Dementia 2019;15:1624–32. 10.1016/j.jalz.2019.08.192.

[47] González HM, Tarraf W, Gouskova N, Gallo LC, Penedo FJ, Davis SM, et al. Neurocognitive function among middle-aged and older hispanic/latinos: Results from the hispanic community health study/study of latinos. Archives of Clinical Neuropsychology 2015;30:68–77. 10.1093/arclin/acu066.

[48] González HM, Tarraf W, Schneiderman N, Fornage M, Vásquez PM, Zeng D, et al. Prevalence and correlates of mild cognitive impairment among diverse Hispanics/Latinos: Study of Latinos-Investigation of Neurocognitive Aging results. Alzheimer’s and Dementia 2019;15:1507–15. 10.1016/j.jalz.2019.08.202.

[49] Zhang Y, Elgart M, Granot-Hershkovitz E, Wang H, Tarraf W, Ramos AR, et al. Genetic associations between sleep traits and cognitive ageing outcomes in the Hispanic Community Health Study/Study of Latinos. EBioMedicine 2023;87. 10.1016/J.EBIOM.2022.104393.

[50] Granot-Hershkovitz E, Spitzer B, Yang Y, Tarraf W, Yu B, Boerwinkle E, et al. Genetic loci of beta-aminoisobutyric acid are associated with aging-related mild cognitive impairment. Transl Psychiatry 2023;13. 10.1038/s41398-023-02437-y.

[51] Albert MS, DeKosky ST, Dickson D, Dubois B, Feldman HH, Fox NC, et al. The diagnosis of mild cognitive impairment due to Alzheimer’s disease: Recommendations from the National Institute on Aging-Alzheimer’s Association workgroups on diagnostic guidelines for Alzheimer’s disease. Alzheimer’s and Dementia 2011;7:270–9. 10.1016/j.jalz.2011.03.008.

[52] Benjaminit Y, Hochberg Y. Controlling the False Discovery Rate: a Practical and Powerful Approach to Multiple Testing. vol. 57. 1995.

[53] Sofer T, Heller R, Bogomolov M, Avery CL, Graff M, North KE, et al. A powerful statistical framework for generalization testing in GWAS, with application to the HCHS/SOL. Genet Epidemiol 2017;41:251–8. 10.1002/gepi.22029.

[54] Chiuve SE, Fung TT, Rimm EB, Hu FB, McCullough ML, Wang M, et al. Alternative dietary indices both strongly predict risk of chronic disease. Journal of Nutrition 2012;142:1009–18. 10.3945/jn.111.157222.

[55] McCullough ML, Maliniak ML, Stevens VL, Carter BD, Hodge RA, Wang Y. Metabolomic markers of healthy dietary patterns in US postmenopausal women. American Journal of Clinical Nutrition 2019;109:1439–51. 10.1093/ajcn/nqy385.

[56] Noerman S, Landberg R. Blood metabolite profiles linking dietary patterns with health—Toward precision nutrition. J Intern Med 2023;293:408–32. 10.1111/joim.13596.

[57] Landberg R, Karra P, Hoobler R, Loftfield E, Huybrechts I, Rattner JI, et al. Dietary biomarkers—an update on their validity and applicability in epidemiological studies. Nutr Rev 2024;82:1260–80. 10.1093/NUTRIT/NUAD119.

[58] Lee G, You HJ, Bajaj JS, Joo SK, Yu J, Park S, et al. Distinct signatures of gut microbiome and metabolites associated with significant fibrosis in non-obese NAFLD. Nature Communications 2020 11:1 2020;11:1–13. 10.1038/s41467-020-18754-5.

[59] Chen B, Bai Y, Tong F, Yan J, Zhang R, Zhong Y, et al. Glycoursodeoxycholic acid regulates bile acids level and alters gut microbiota and glycolipid metabolism to attenuate diabetes. Gut Microbes 2023;15. 10.1080/19490976.2023.2192155.

[60] https://phytohub.eu/entries/PHUB001078 2025.

[61] Teixeira J, Gaspar A, Garrido EM, Garrido J, Borges F. Hydroxycinnamic acid antioxidants: An electrochemical overview. Biomed Res Int 2013;2013. 10.1155/2013/251754.

[62] Hu J, Chen J, Xu X, Hou Q, Ren J, Yan X. Gut microbiota-derived 3-phenylpropionic acid promotes intestinal epithelial barrier function via AhR signaling. Microbiome 2023;11. 10.1186/s40168-023-01551-9.

[63] Li P, Feng X, Ma Z, Yuan Y, Jiang H, Xu G, et al. Microbiota-derived 3-phenylpropionic acid promotes myotube hypertrophy by Foxo3/NAD+ signaling pathway. Cell Biosci 2024;14. 10.1186/s13578-024-01244-2.

[64] Zuraikat FM, Wood RA, Barragán R, St-Onge MP. Sleep and Diet: Mounting Evidence of a Cyclical Relationship. Annu Rev Nutr 2021;41:309–32. 10.1146/annurev-nutr-120420-021719.

[65] Liu Y, Tabung FK, Stampfer MJ, Redline S, Huang T. Overall diet quality and proinflammatory diet in relation to risk of obstructive sleep apnea in 3 prospective US cohorts. American Journal of Clinical Nutrition 2022;116:1738–47. 10.1093/ajcn/nqac257.

[66] Lieberman HR, Agarwal S, Caldwell JA, Fulgoni VL. Demographics, sleep, and daily patterns of caffeine intake of shift workers in a nationally representative sample of the US adult population. Sleep 2020;43. 10.1093/sleep/zsz240.

[67] Nea FM, Kearney J, Livingstone MBE, Pourshahidi LK, Corish CA. Dietary and lifestyle habits and the associated health risks in shift workers. Nutr Res Rev 2015;28:143–66. 10.1017/S095442241500013X.

[68] Aragozzini F, Ferrari A, Pacini N, Gualandris R. NOTES Indole-3-Lactic Acid as a Tryptophan Metabolite Produced by Bifidobacterium spp. vol. 38. 1979.

[69] Chen Y, Fang JY. The role of colonic microbiota amino acid metabolism in gut health regulation. Cell Insight 2025;4. 10.1016/j.cellin.2025.100227.

[70] Hu Y, Li J, Wang B, Zhu L, Li Y, Ivey KL, et al. Interplay between diet, circulating indolepropionate concentrations and cardiometabolic health in US populations. Gut 2023;72:2260–71. 10.1136/gutjnl-2023-330410.

[71] Menni C, Zhu J, Le Roy CI, Mompeo O, Young K, Rebholz CM, et al. Serum metabolites reflecting gut microbiome alpha diversity predict type 2 diabetes. Gut Microbes 2020;11:1632–42. 10.1080/19490976.2020.1778261.

[72] Peters BA, Qi Q, Usyk M, Daviglus ML, Cai J, Franceschini N, et al. Association of the gut microbiome with kidney function and damage in the Hispanic Community Health Study/Study of Latinos (HCHS/SOL). Gut Microbes 2023;15. 10.1080/19490976.2023.2186685.

[73] Pallister T, Jennings A, Mohney RP, Yarand D, Mangino M, Cassidy A, et al. Characterizing blood metabolomics profiles associated with self-reported food intakes in female twins. PLoS One 2016;11. 10.1371/journal.pone.0158568.

[74] Naja K, Anwardeen N, Al-Shafai M, Elrayess MA. Indoleacetylglutamine Pathway Is a Potential Biomarker for Cardiovascular Diseases. Biomolecules 2025;15. 10.3390/biom15030377.

[75] Lustgarten MS, Fielding RA. Metabolites Associated With Circulating Interleukin-6 in Older Adults. J Gerontol A Biol Sci Med Sci 2017;72:1277–83. 10.1093/gerona/glw039.

[76] Serger E, Luengo-Gutierrez L, Chadwick JS, Kong G, Zhou L, Crawford G, et al. The gut metabolite indole-3 propionate promotes nerve regeneration and repair. Nature 2022;607:585–92. 10.1038/s41586-022-04884-x.

[77] Xiao HW, Cui M, Li Y, Dong JL, Zhang SQ, Zhu CC, et al. Gut microbiota-derived indole 3-propionic acid protects against radiation toxicity via retaining acyl-CoA-binding protein. Microbiome 2020;8. 10.1186/S40168-020-00845-6,.

[78] Poeggeler B, Pappolla MA, Hardeland R, Rassoulpour A, Hodgkins PS, Guidetti P, et al. Indole-3-propionate: A potent hydroxyl radical scavenger in rat brain. Brain Res 1999;815:382–8. 10.1016/S0006-8993(98)01027-0,.

[79] Xiao HW, Cui M, Li Y, Dong JL, Zhang SQ, Zhu CC, et al. Gut microbiota-derived indole 3-propionic acid protects against radiation toxicity via retaining acyl-CoA-binding protein. Microbiome 2020;8. 10.1186/S40168-020-00845-6,.

[80] Sweetman A, Lack L, McEvoy RD, Smith S, Eckert DJ, Osman A, et al. Bi-directional relationships between co-morbid insomnia and sleep apnea (COMISA). Sleep Med Rev 2021;60. 10.1016/j.smrv.2021.101519.

[81] Shahu A, Chung J, Tarraf W, Ramos AR, González HM, Redline S, et al. Method comparison and estimation of causal effects of insomnia on health outcomes in a survey sampled population. Sci Rep 2023;13. 10.1038/s41598-023-36927-2.

[82] He S, Granot-Hershkovitz E, Zhang Y, Bressler J, Tarraf W, Yu B, et al. Blood metabolites predicting mild cognitive impairment in the study of Latinos-investigation of neurocognitive aging (HCHS/SOL). Alzheimers Dement (Amst) 2022;14. 10.1002/DAD2.12259.

